# Should we screen less frequently for chlamydia and gonorrhoea in gay and bisexual men who have sex with men? Findings from a global crowdsourcing exercise with experts

**DOI:** 10.1101/2025.10.15.25337958

**Authors:** Teralynn Ludwick, Ethan T Cardwell, Tin D. Vo, Lauren Ware, Phoebe Quinn, Eric PF Chow, Daniel Grace, Jane S Hocking, Fabian YS Kong

## Abstract

**Objectives:** Many countries recommend 3-monthly chlamydia/gonorrhoea screening for men-who-have-sex-with-men (MSM). New evidence about the limited impact of frequent, asymptomatic gonorrhoea/chlamydia screening on population prevalence, coupled with concerns about overburdened health services and antimicrobial resistance (from over-treatment), calls into question current approaches to asymptomatic screening. We explored sexual health professionals/experts’ arguments in favour/against reducing asymptomatic screening using Polis (www.Pol.is), an online, crowdsourcing tool for understanding what large groups think.

**Methods:** Recruited via global peak bodies/networks, 99 sexual health professionals/experts (43.4.% clinicians, 35.4% researchers) primarily from Oceania (41.4%), UK/Europe (29.4%) and North America (22.2%) submitted 83 statements in favour/against reduced screening for men-who-have-sex-with-men (e.g. ‘Bisexual men who don’t test regularly risk putting women at risk’). Participants voted on submitted statements (agree/disagree/pass). We considered statements with _≥_80 agreement as strong support, 70-79% moderate support, _≤_69% mixed support. We used content analysis to group clusters of related statements, and examined associations between participant demographics and votes for/against.

**Results:** There was ‘mixed support’ for statements on :1) the impact of screening in reducing prevalence; 2) whether asymptomatic infections pose clinical harm/necessitate treatment; and risk of antimicrobial resistance. Statements advocating for 6-monthly screening received ‘moderate support’, with arguments centering on resource use. Participants ‘strongly supported’ the need for community engagement and maintaining frequent HIV/syphilis screening. UK/Europe participants were more likely to favour reduced chlamydia/gonorrhoea screening.

**Conclusions:** While there were mixed opinions about relative utility, risks, and harms of reducing chlamydia/gonorrhoea screening for MSM, arguments relating to resource use may provide common ground for policy changes.

## Introduction

Asymptomatic screening is an important component of STI control strategies in many countries, including for gay, bisexual and other men who have sex with men (GBMSM) who are disproportionately affected by sexually transmitted infections (STIs).^1,2^ Many countries recommend 3-monthly HIV/STI screening for sexually active GBMSM to reduce sequelae and transmission,^3,4^ and for routine monitoring of those on HIV pre-exposure prophylaxis (PrEP).

Frequent screening is championed by public health authorities, HIV/AIDS service providers, and GBMSM community organisations as a tenet of good sexual citizenship.^5^ While there is strong evidence that 3-monthly screening for HIV and syphilis (etiological agent *Treponema pallidum*) reduces transmission and morbidity,^6,7^ 3-monthly screening for *Chlamydia trachomatis* (CT) and *Neisseria gonorrhoeae* (NG) appears to have little impact on the prevalence of these STIs.^8,9^ Studies show that the higher rates and transmission of NG among GBMSM are due to dense sexual networks, and consequently frequent screening has little impact on prevalence in this population as the high-equilibrium prevalence is sustained through frequent reinfections within this network.^10^

Consequently, some experts are questioning whether the harms of 3-monthly CT/NG screening outweigh benefits for GBMSM.^11-13^ About 80% of detected CT and NG cases are asymptomatic, pose little harm in cis-males, and often clear spontaneously 6-to-16 weeks without treatment.^14,15^ However, the high incidence of asymptomatic CT/NG detected by 3-monthly screening has led to significant increases in antibiotic prescribing,^16^ contributing to drug-resistant NG (a critical health threat designated by the World Health Organisation) and broader antimicrobial resistance (AMR) at the population level.^17^ Due to the limited availability of new classes of antibiotics (including broad-spectrum), it is important to consider their judicious use within the context of CT/NG screening and management. Further, there are concerns about the significant costs incurred by health systems and individuals related to frequent screening.^18^

While countries like the Netherlands and Belgium have moved to reduce and/or eliminate asymptomatic CT/NG screening,^19,20^ global support for doing so is contested and represents a major departure from the well-established ‘test and treat’ paradigm. Concerns about reducing screening frequency include reduced opportunities for health system engagement and potentially negative impacts to HIV and syphilis screening (for which missed and/or delayed detection has serious consequences). Qualitative research has shown that GBMSM support treatment of asymptomatic CT/NG infections and value the sense of safety and control over sexual health afforded by 3-monthly screening.^21^

There is a need for greater global consensus on how to manage asymptomatic CT/NG screening and how to balance different public health trade-offs.^13,22,23^ This study used a crowdsourcing platform called *Polis* (www.Pol.is) to solicit global views on the acceptability of reducing screening frequency for asymptomatic CT and NG in GBMSM. Through crowdsourcing global expert opinion, this study aimed to: (1) identify areas of (dis)agreement within the sexual health community; (2) raise areas for further education to support a more informed debate; and contribute to future policy directions.

## Methods

To canvas global views on the acceptability of reduced asymptomatic screening for CT/NG in GBMSM, we used *Polis* (www.Pol.is), a data collection tool known as a ‘wiki survey’ which incorporates automated, real-time analysis. In response to an open-ended prompt, *Polis* enables participants to submit short responses in their own words, which are then appraised by other participants via collective voting. *Polis* has been used as a consultation tool in democratic decision-making processes and academic research.^24-26^

Professionals working in the STI field (including clinicians, researchers, policymakers, and community representatives) were invited to respond to the question: “Should we continue to conduct regular asymptomatic screening for chlamydia and gonorrhoea among GBMSM?” To spur a nuanced discussion and encourage participants to consider the harms, benefits, and implementation considerations around changing guidelines, we provided three sub-questions:

1. Given concerns about rising antibiotic resistance, should guidelines around frequency of asymptomatic screening for CT and NG for GBMSM change?
2. How should we manage asymptomatic cases of chlamydia and gonorrhoea?
3. What would be the unintended consequences of changes in screening guidelines for chlamydia and gonorrhoea (e.g. on other STIs, for GBMSM or other populations)? What should be done to mitigate these consequences?

To initiate the conversation, we provided eight ‘seed statements’ covering a wide range of potential opinions and concerns across the 3 sub-questions (Appendix 1). The seed statements were vetted and piloted with a small expert group. Participants were asked to vote on the seed statements (agree, disagree, or pass), and add their own opinion statements if they wished.

Statements are presented in a semi-random order based on *Polis*’ prioritisation algorithms which help to manage large volumes of statements.^24^ Participants’ statements were accepted without modifications, but rejected if off-topic, non-sensical, or very similar to existing statements without adding additional nuance (Appendix 2).

### Recruitment

We used networks and snowballing to recruit individuals from diverse occupations within the STI field. We invited members of: (1) International Union against STIs (IUSTI) – the field’s apex organization with hundreds of members; (2) conference organizing committees for key upcoming international STI conferences; (3) World Health Organization Collaborating Centre for STIs; (4) US Centers for Disease Control National Network of STD Clinical Prevention Training Centres; (5) Australasian Society for HIV, Viral Hepatitis and Sexual Health Medicine (ASHM); and, (6) Editors of STI journals. We encouraged all those contacted to share the invitation widely.

The invitation contained a weblink to a registration survey (managed via Qualtrics) in which individuals recorded their interest, provided consent, and answered questions about their background and expertise (Appendix 4). Once participants submitted the form, they received an anonymized weblink to access the *Polis* conversation. As the conversation evolved in real-time, participants received weekly reminders over a 3-week period in August 2024 to respond to new opinion statements submitted by participants. Participants could opt to engage as little or as often as they liked. Thus, the numbers of votes vary across statements, tending to be lower on comments submitted nearer to the closing date (indicated by higher statement ID number) (Appendix 3).

### Analysis

*Polis* software generates an interactive report which displays all approved statements and the number of votes (agreed, disagreed, or passed) for each statement (Figure 1).

**Figure 1:**
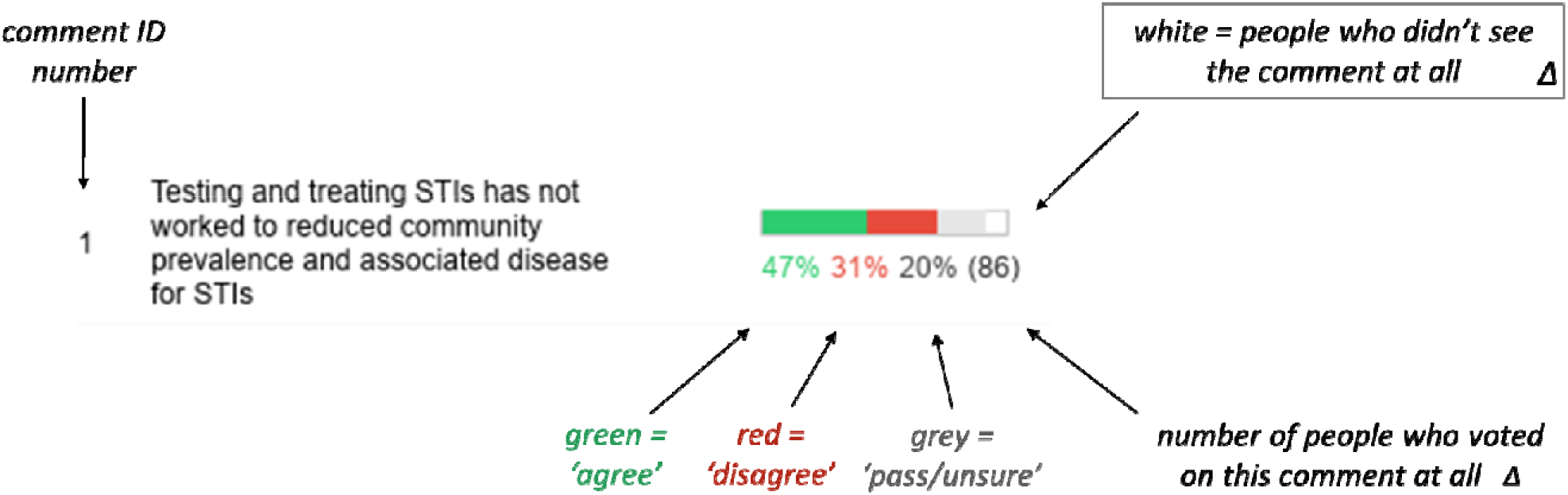
Explanation of *Polis* Bar Chart.

Using content analysis, we first categorised the statements according to our three previously identified sub-questions. We then used an inductive process to develop additional categories. Statements could be coded to multiple categories, where relevant.

Within each category, we grouped statements together which addressed similar views in order to understand voting patterns and to interpret relative support for clusters of related ideas. We considered statements with _≥_80% agreement as ‘strong’ support, 70-79% as ‘moderate’ support and anything below as ‘mixed’ support. In drawing inferences, we took into account the percentage of ‘disagree’ and ‘pass/uncertain’ votes, as well as the total number of votes on a given statement.

Using principal component analysis and k-means clustering, *Polis* generates ‘opinion groups’ based on participants who voted similarly to each other across a range of statements, and differently from other groups.^24^ In our project, Polis generated two opinion groups. To assess differences in participants categorized in these two groups (A and B), we conducted Fisher’s exact tests to compare their demoraphic characteristics.

Study approval was granted by the University of Melbourne Human Research Ethics Committee (ID_2024-28374-58750-7).

## Results

### Participant characteristics and level of engagement

Ninety-nine experts and professionals participated by voting on and/or submitting opinion statements outlining considerations in favour of or against frequent screening. Over a 3-week period, 91 opinion statements were submitted (including the original 8 seed statements). On average, each participant voted on 75.8% (69/91) of all statements, ranging from 1 to 85 votes cast per participant.

Participants were mainly clinicians (43·4%) and public health researchers (27·3%), primarily from Australia/New Zealand (41·4%), the UK/Europe (29·3%), and North America (22·2%). Participants most commonly worked in public healthcare (32.3%), universities/research institutes (31·3%), or government (21·2%). Among participants, 61·6% identified as women and 41·4% as part of the LGBTIQ+ community. A larger percentage (81.8%) of participants self-rated their knowledge of CT/NG treatment as ‘expert or good’, compared to only 68·7% on their knowledge of AMR. Overall 0·7% perceived AMR as a threat in their own country (Table 1).

**Table 1:** Participant characteristics, knowledge, and views.

|  |  | n=99 | % |
| --- | --- | --- | --- |
| **Gender** | Woman | 61 | 61.6% |
|  | Man | 35 | 35.4% |
|  | Non-binary | 3 | 3.0% |
| **LGBTIQ+<sup>1</sup>** | Yes | 41 | 41.4% |
| **Profession** | GP | 2 | 2.0% |
|  | Nurse/allied health | 18 | 18.2% |
|  | Medical infectious disease specialist | 6 | 6.1% |
|  | Sexual health physician | 17 | 17.2% |
|  | Education/social work/counselling/Health promotion | 7 | 7.1% |
|  | Diagnostics/Laboratory | 5 | 5.1% |
|  | Policy | 3 | 3.0% |
|  | Other | 6 | 6.1% |
|  | Research - public health/epidemiology | 27 | 27.3% |
|  | Research - other | 8 | 8.1% |
| **Primary work setting** | Government | 21 | 21.2% |
|  | NGO | 11 | 11.1% |
|  | Private healthcare | 2 | 2.0% |
|  | Public Healthcare | 32 | 32.3% |
|  | University/Research Institute | 31 | 31.3% |
|  | Other | 2 | 2.0% |
| **Region** | Africa/Asia/Latin America | 7 | 7.1% |
|  | North America | 22 | 22.2% |
|  | Oceania | 41 | 41.4% |
|  | UK and Europe | 29 | 29.3% |
| **Knowledge regarding management of NG<sup>2</sup> and CT<sup>3</sup>** | Expert | 38 | 38.4% |
|  | Good | 43 | 43.4% |
|  | Moderate | 15 | 15.2% |
|  | Limited | 2 | 2.0% |
|  | None | 1 | 1.0% |
| **Knowledge of AMR<sup>4</sup>** | Expert | 25 | 25.3% |
|  | Good | 43 | 43.4% |
|  | Moderate | 8 | 8.1% |
|  | Limited | 22 | 22.2% |
|  | None | 1 | 1.0% |
| **AMR is a global public health threat** | Strongly agree | 83 | 83.8% |
|  | Generally agree | 11 | 11.1% |
|  | Neutral | 2 | 2.0% |
|  | Generally disagree | 0 | 0.0% |
|  | Strongly disagree | 3 | 3.0% |
| **AMR is a NOT a threat in my country** | Strongly agree | 8 | 8.1% |
|  | Generally, agree | 9 | 9.1% |
|  | Neutral | 10 | 10.1% |
|  | Generally disagree | 23 | 23.2% |
|  | Strongly disagree | 47 | 47.5% |
|  | Missing | 2 | 2.0% |

1. LGBTQIA+ refers to identifying as a member of the lesbian, gay, bisexual, transgender, queer, intersex and asexual community.
2. NG refers to Neisseria gonorrhoeae
3. CT refers to chlamydia trachomatis
4. AMR refers to antimicrobial resistance

### 2. Content analysis of opinion statements and voting patterns

Four main subjects of discussion merged: (1) the role of screening in reducing community prevalence; (2) considerations on the frequency of screening; (3) management and treatment; and, (4) positive externalities and managing unintended consequences. Tables 2.1-2.4 include illustrative examples of statements with stronger or lower levels of support to reflect the overall voting patterns. See Appendix 3 for full list of statements.

**Table 2.1.**
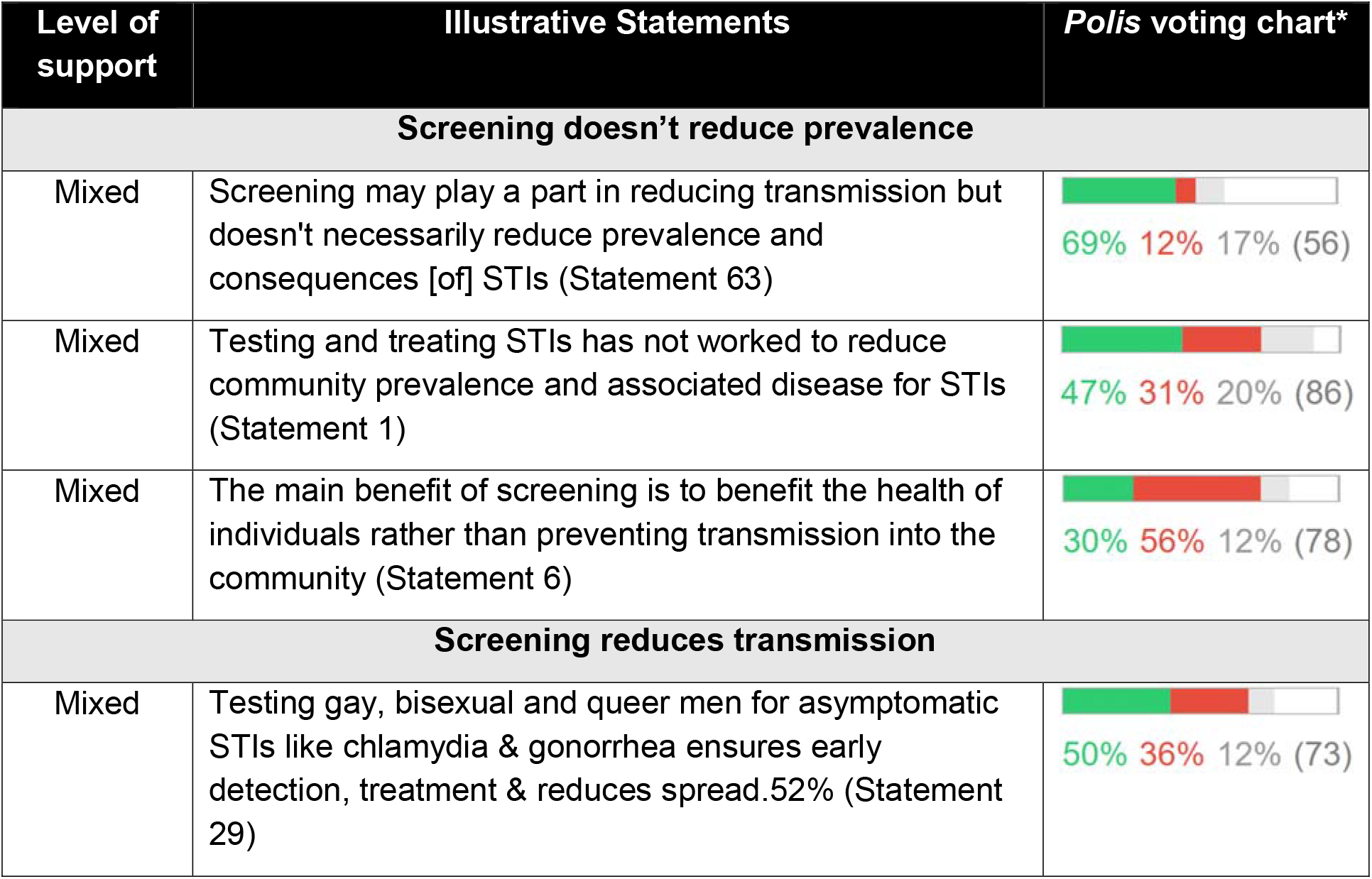
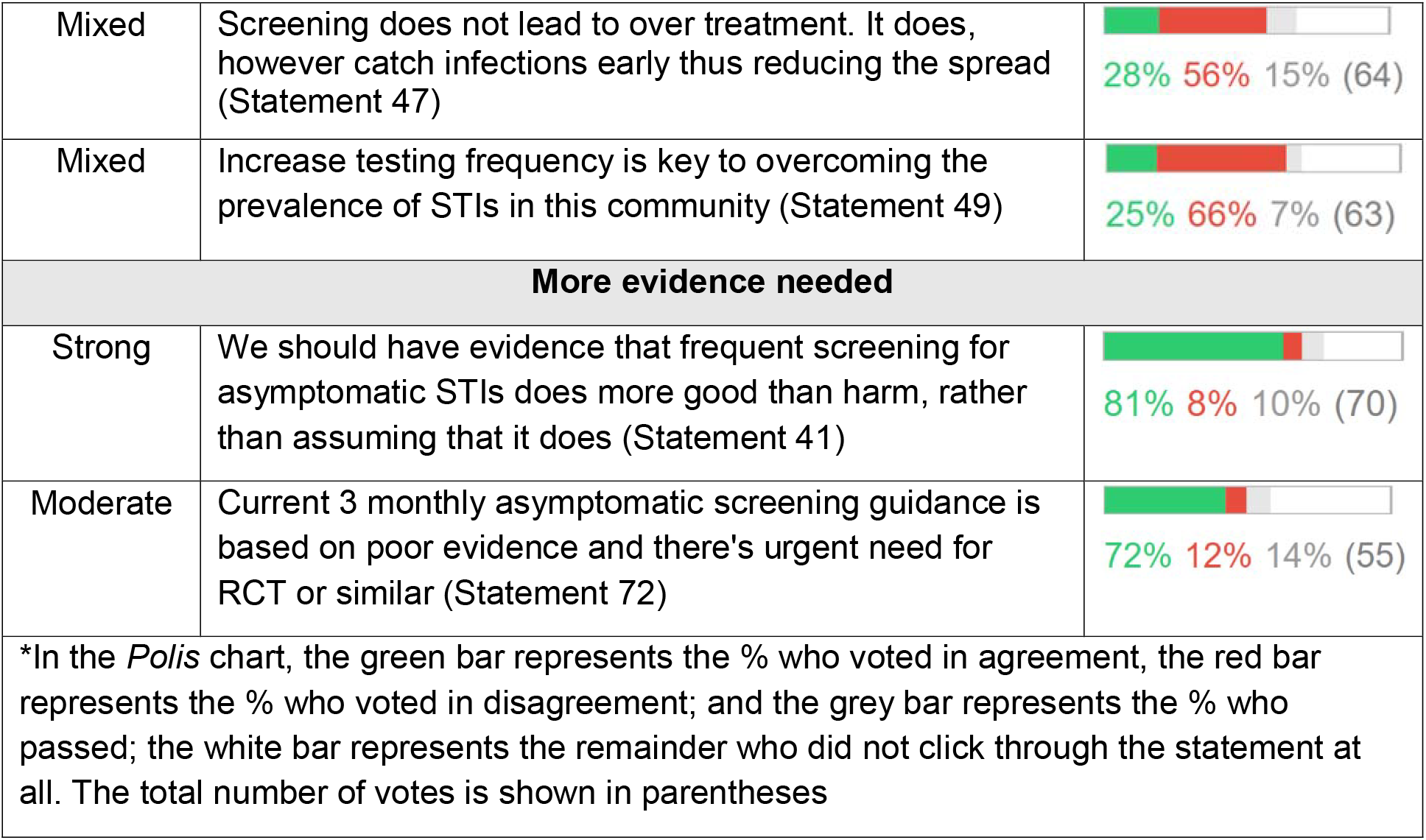
Illustrative statements – Opinions on the role of screening in reducing prevalence.

| Level of support | Illustrative Statements | Polis voting chart* |
| --- | --- | --- |
| **Screening doesn’t reduce prevalence** |  |  |
| Mixed | Screening may play a part in reducing transmission but doesn’t necessarily reduce prevalence and consequences [of] STIs (Statement 63) | 69% 12% 17% (56) |
| Mixed | Testing and treating STIs has not worked to reduce community prevalence and associated disease for STIs (Statement 1) | 47% 31% 20% (86) |
| Mixed | The main benefit of screening is to benefit the health of individuals rather than preventing transmission into the community (Statement 6) | 30% 56% 12% (78) |
| **Screening reduces transmission** |  |  |
| Mixed | Testing gay, bisexual and queer men for asymptomatic STIs like chlamydia & gonorrhea ensures early detection, treatment & reduces spread.52% (Statement 29) | 50% 36% 12% (73) |
| Mixed | Screening does not lead to over treatment. It does, however catch infections early thus reducing the spread (Statement 47) | 28% 56% 15% (64) |
| Mixed | Increase testing frequency is key to overcoming the prevalence of STIs in this community (Statement 49) | 25% 66% 7% (63) |
| **More evidence needed** |  |  |
| Strong | We should have evidence that frequent screening for asymptomatic STIs does more good than harm, rather than assuming that it does (Statement 41) | 81% 8% 10% (70) |
| Moderate | Current 3 monthly asymptomatic screening guidance is based on poor evidence and there's urgent need for RCT or similar (Statement 72) | 72% 12% 14% (55) |
| *In the *Polis* chart, the green bar represents the % who voted in agreement, the red bar represents the % who voted in disagreement; and the grey bar represents the % who passed; the white bar represents the remainder who did not click through the statement at all. The total number of votes is shown in parentheses |  |  |

**Table 2.2.**
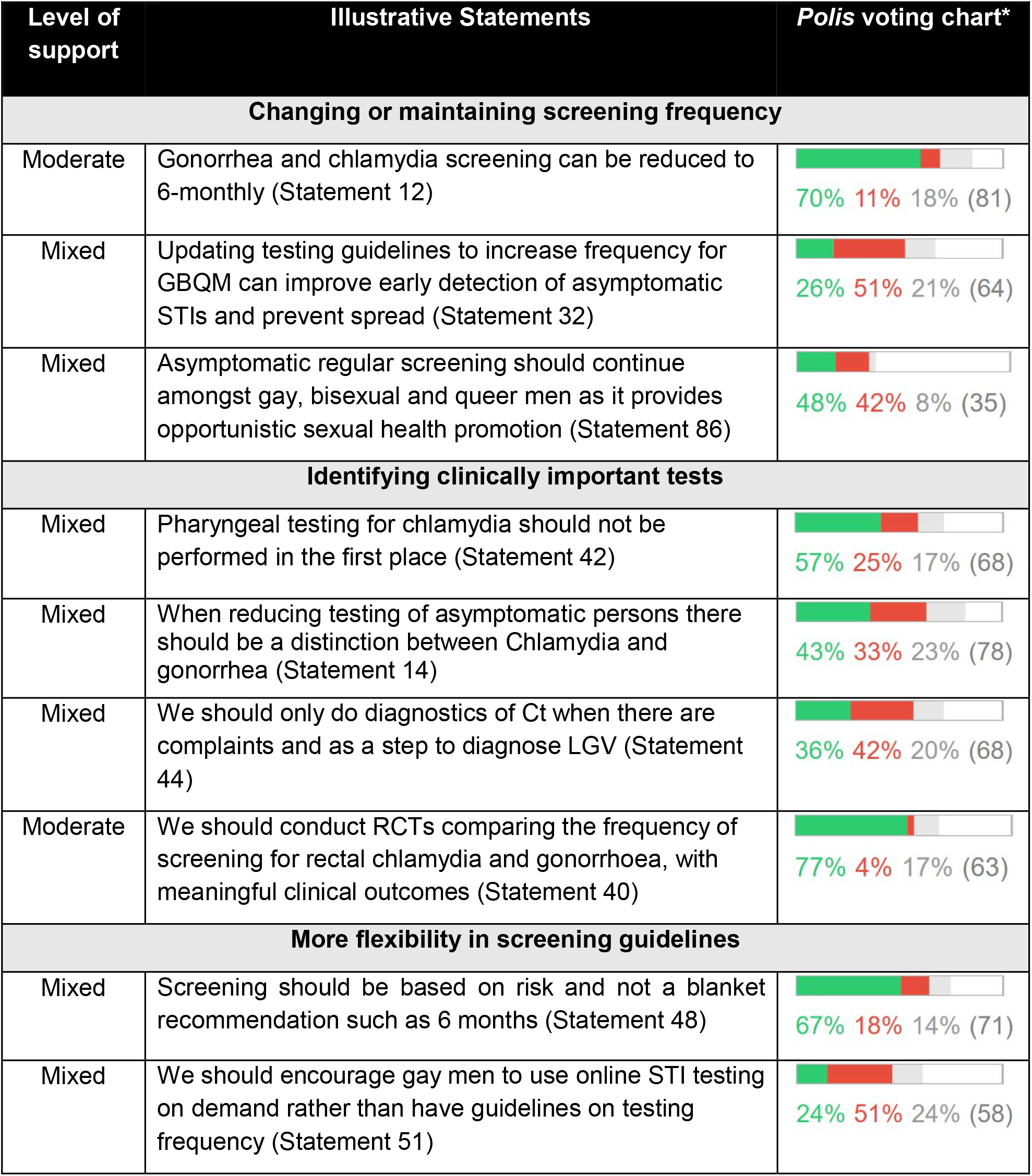
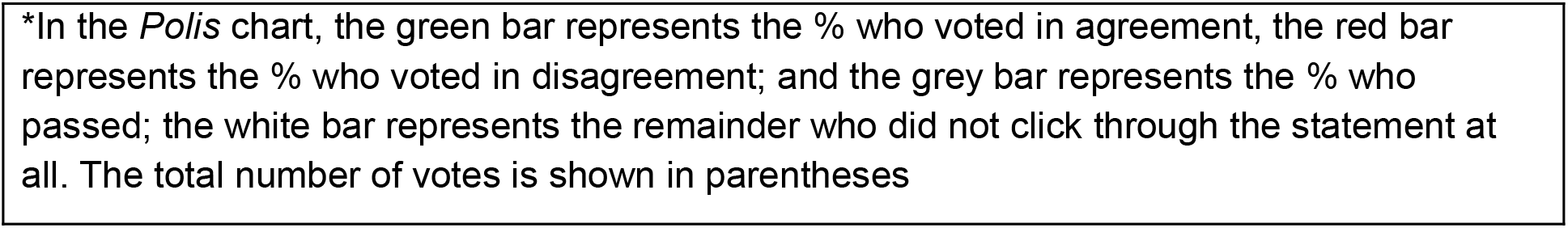
Illustrative statements – Opinions and considerations on screening frequency for CT and NG in GBMSM.

| Level of support | Illustrative Statements | Polis voting chart* |
| --- | --- | --- |
| **Changing or maintaining screening frequency** |  |  |
| Moderate | Gonorrhea and chlamydia screening can be reduced to 6-monthly (Statement 12) | 70% 11% 18% (81) |
| Mixed | Updating testing guidelines to increase frequency for GBQM can improve early detection of asymptomatic STIs and prevent spread (Statement 32) | 26% 51% 21% (64) |
| Mixed | Asymptomatic regular screening should continue amongst gay, bisexual and queer men as it provides opportunistic sexual health promotion (Statement 86) | 48% 42% 8% (35) |
| **Identifying clinically important tests** |  |  |
| Mixed | Pharyngeal testing for chlamydia should not be performed in the first place (Statement 42) | 57% 25% 17% (68) |
| Mixed | When reducing testing of asymptomatic persons there should be a distinction between Chlamydia and gonorrhea (Statement 14) | 43% 33% 23% (78) |
| Mixed | We should only do diagnostics of Ct when there are complaints and as a step to diagnose LGV (Statement 44) | 36% 42% 20% (68) |
| Moderate | We should conduct RCTs comparing the frequency of screening for rectal chlamydia and gonorrhoea, with meaningful clinical outcomes (Statement 40) | 77% 4% 17% (63) |
| **More flexibility in screening guidelines** |  |  |
| Mixed | Screening should be based on risk and not a blanket recommendation such as 6 months (Statement 48) | 67% 18% 14% (71) |
| Mixed | We should encourage gay men to use online STI testing on demand rather than have guidelines on testing frequency (Statement 51) | 24% 51% 24% (58) |

\*In the *Polis* chart, the green bar represents the % who voted in agreement, the red bar represents the % who voted in disagreement; and the grey bar represents the % who passed; the white bar represents the remainder who did not click through the statement at all. The total number of votes is shown in parentheses

**Table 2.3:**
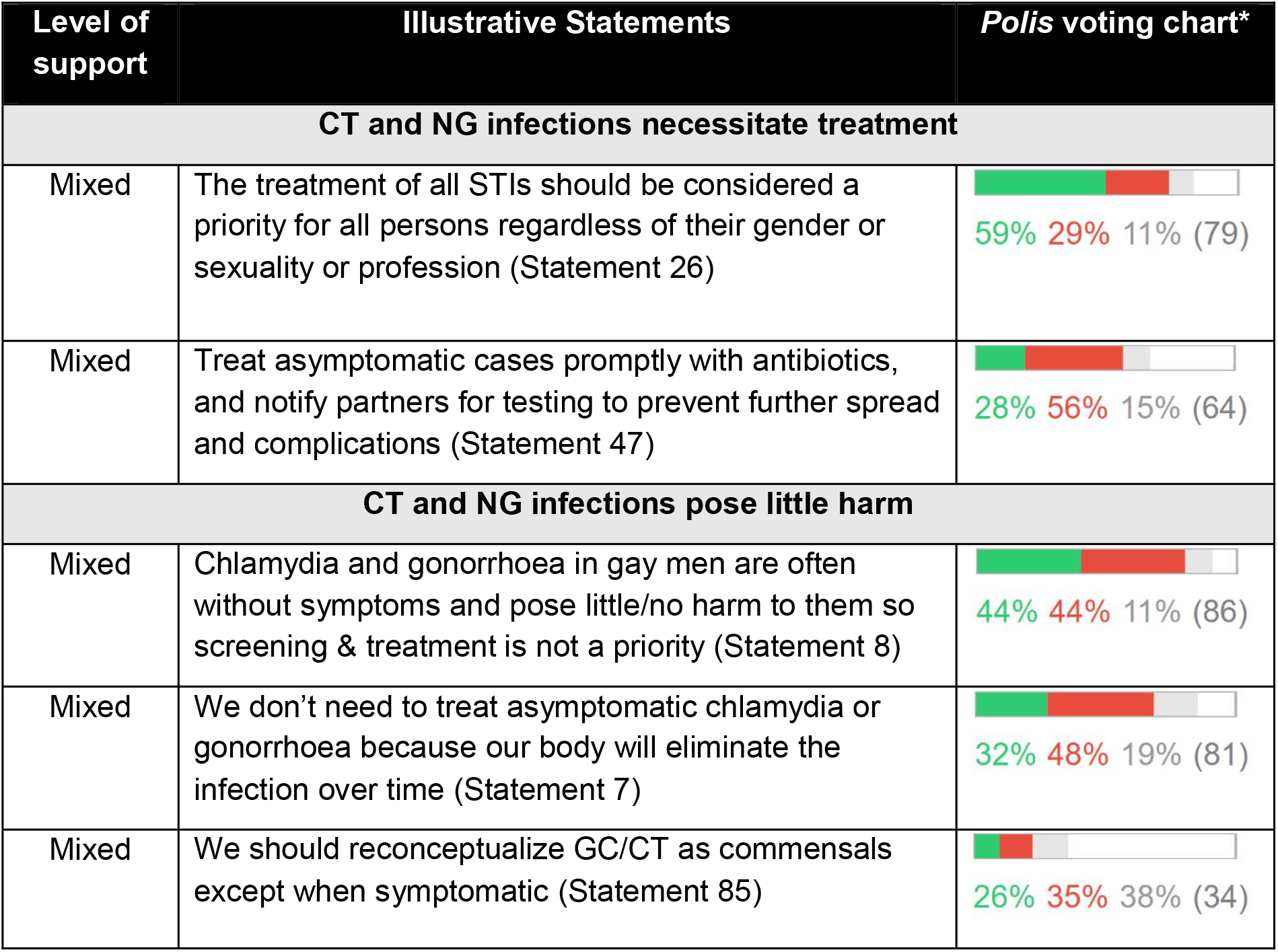
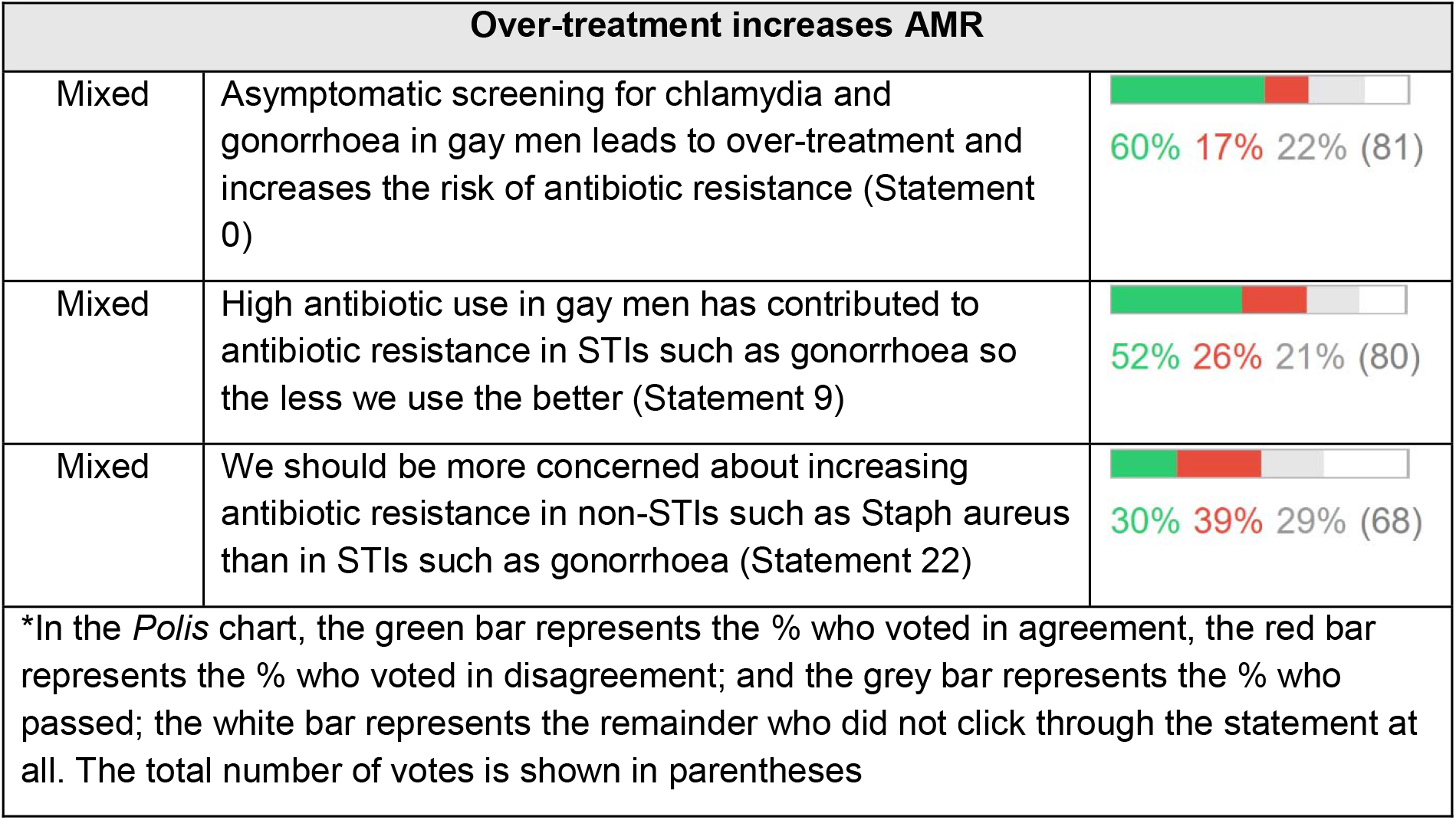
Illustrative statements - Opinions on the management and treatment of asymptomatic cases of chlamydia and gonorrhoea in GBMSM.

| Level of support | Illustrative Statements | *Polis* voting chart* |
| --- | --- | --- |
| **CT and NG infections necessitate treatment** |  |  |
| Mixed | The treatment of all STIs should be considered a priority for all persons regardless of their gender or sexuality or profession (Statement 26) | 59% 29% 11% (79) |
| Mixed | Treat asymptomatic cases promptly with antibiotics, and notify partners for testing to prevent further spread and complications (Statement 47) | 28% 56% 15% (64) |
| **CT and NG infections pose little harm** |  |  |
| Mixed | Chlamydia and gonorrhoea in gay men are often without symptoms and pose little/no harm to them so screening & treatment is not a priority (Statement 8) | 44% 44% 11% (86) |
| Mixed | We don't need to treat asymptomatic chlamydia or gonorrhoea because our body will eliminate the infection over time (Statement 7) | 32% 48% 19% (81) |
| Mixed | We should reconceptualize GC/CT as commensals except when symptomatic (Statement 85) | 26% 35% 38% (34) |

| Over-treatment increases AMR |  |  |
| --- | --- | --- |
| Mixed | Asymptomatic screening for chlamydia and gonorrhoea in gay men leads to over-treatment and increases the risk of antibiotic resistance (Statement 0) | 60% 17% 22% (81) |
| Mixed | High antibiotic use in gay men has contributed to antibiotic resistance in STIs such as gonorrhoea so the less we use the better (Statement 9) | 52% 26% 21% (80) |
| Mixed | We should be more concerned about increasing antibiotic resistance in non-STIs such as Staph aureus than in STIs such as gonorrhoea (Statement 22) | 30% 39% 29% (68) |
| *In the *Polis* chart, the green bar represents the % who voted in agreement, the red bar represents the % who voted in disagreement; and the grey bar represents the % who passed; the white bar represents the remainder who did not click through the statement at all. The total number of votes is shown in parentheses |  |  |

**Table 2.4.**
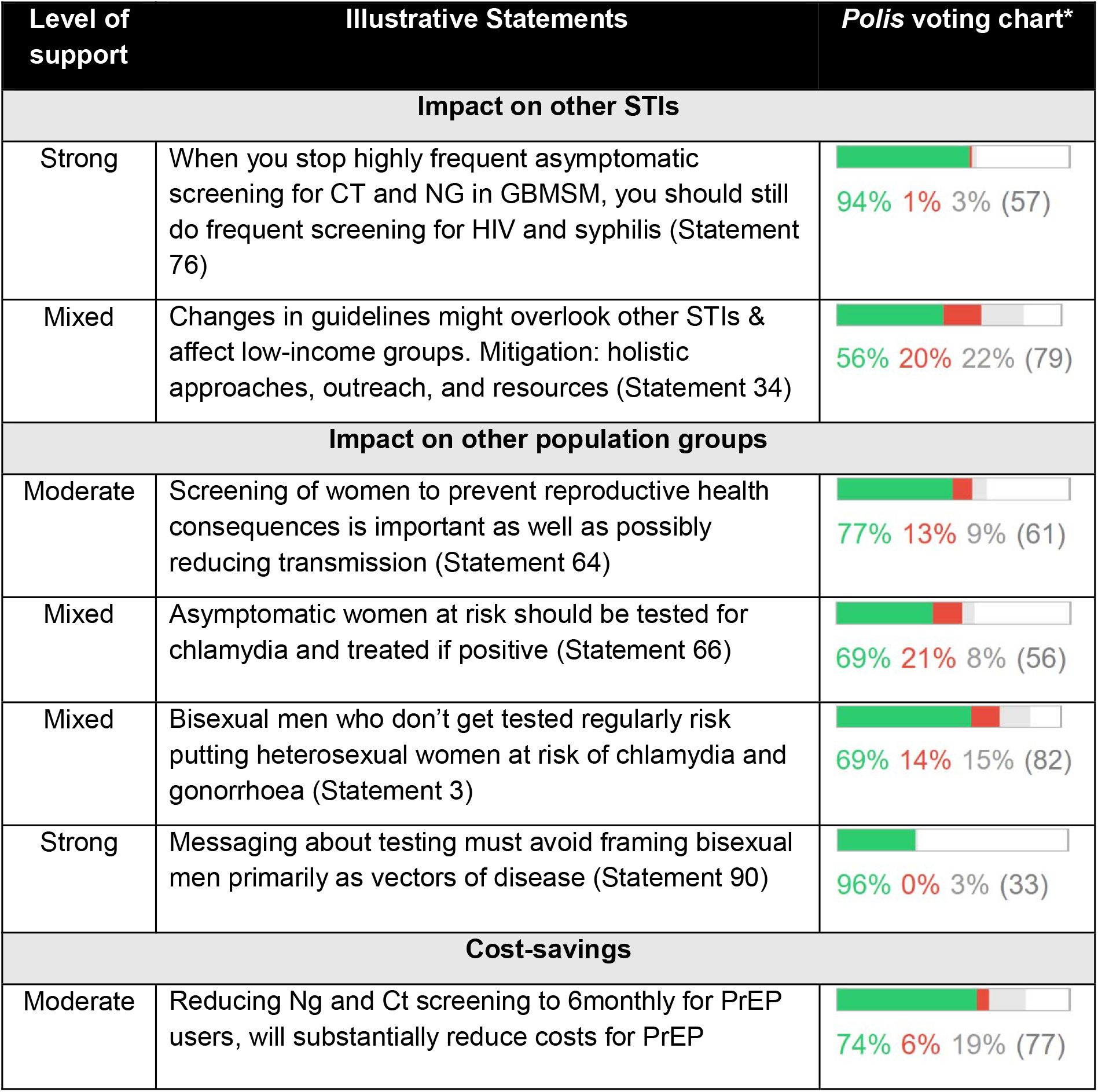
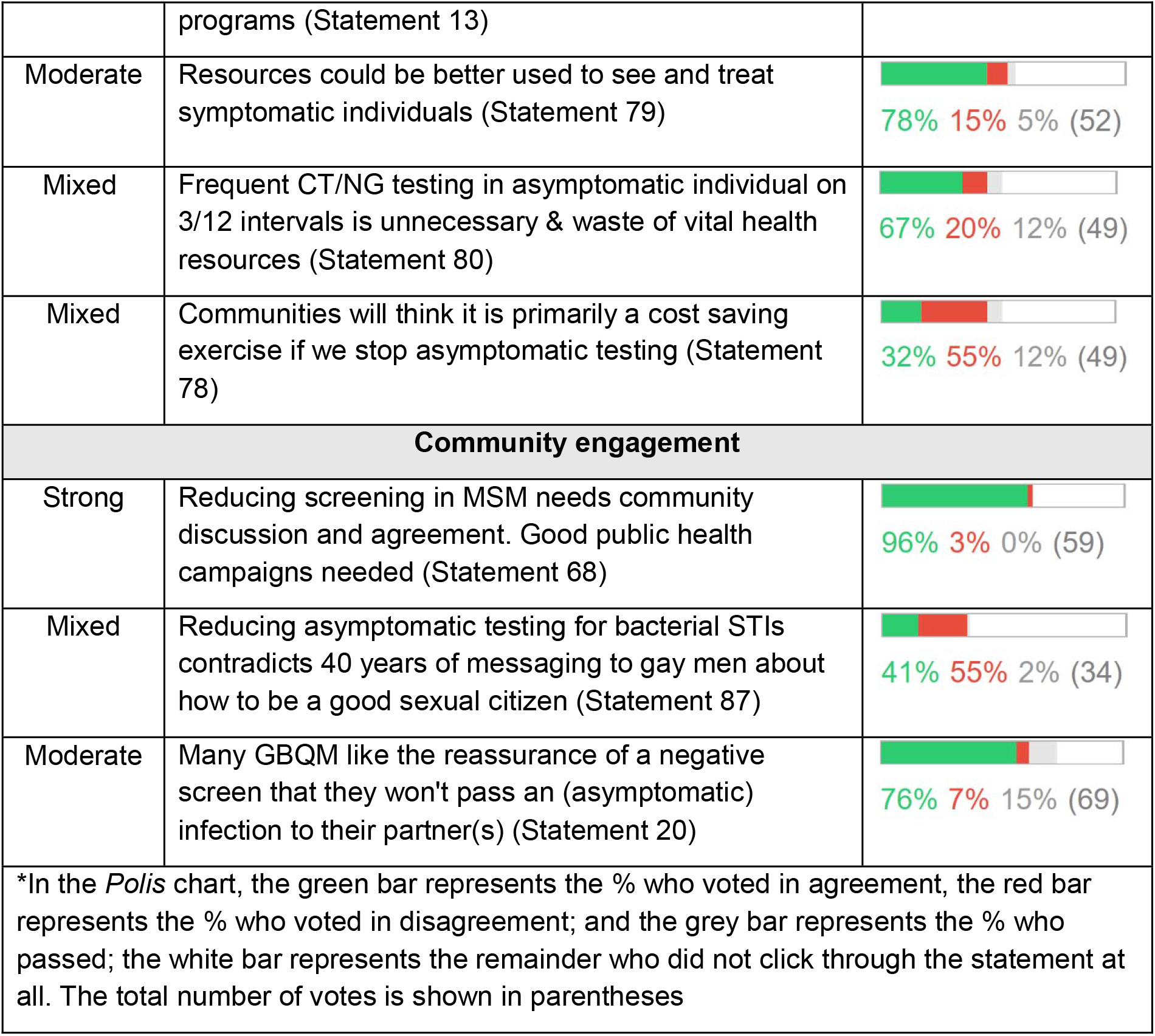
Illustrative statements - Positive externalities and managing unintended consequences of reduced CT and NG screening frequency.

#### 2.1 Role of screening in reducing community prevalence

There was mixed support for statements conveying the inutility of screening in reducing prevalence (Statement_1/63). Similarly, statements which affirmed the value of the traditional ‘test and treat’ paradigm in ‘reducing spread’ received mixed support (Statements_29/31/47/54). Lack of consensus in this area was further compounded by mixed opinion about whether the main benefit of screening is to reduce population prevalence or to support individual health (Statements_6).

With no clear consensus on the impact of screening on community prevalence, there was moderate to strong support for several statements calling for more evidence, particularly for randomized controlled trials to inform guidelines (Statements_72/75/82) and to demonstrate benefits over harms (Statement_41). In contrast, the statement opining that reductions in STI screening frequency will ‘compromise our understanding of community STI prevalence’ received mixed support (Statement_38). Table 2.1 presents illustrative statements.

#### 2.2 Considerations for screening frequency

Two statements explicitly proposed reducing screening frequency to 6-monthly, both of which had moderate support (Statements_12/13). A large number of statements underscored reasons for reducing screening frequency, including being a waste of resources (elaborated in section 2.4) and being clinically unnecessary. However, there were mixed views on what aspects of screening were clinically (un)important, including whether testing should focus on certain anatomical sites, strains, or specific (heterosexual) populations (Statements_14/15/39/40/42/44/46/70/74). There was nothing close to consensus on whether to:

- eliminate pharyngeal screening for CT;
- limit CT screening to Lymphogranuloma venereum (LGV-type) only;
- limit CT screening for diagnosing symptomatic cases in women and partners;
- test rectal sites over other anatomical sites;
- differentiate screening guidelines for NG versus CT;
- screen for NG based on the correlation between NG reinfection with HIV acquisition;
- screen for CT/NG if on HIV PrEP.

In contrast, two statements called for increasing screening frequency as a means of reducing prevalence, but only a minority agreed (Statements_32/49). There was mixed support for a statement in favour of regular screening to provide opportunistic sexual health promotion (Statement_86). Aligning with the high level of uncertainty on many clinical aspects, there was moderate support for new studies to assess screening at different anatomical sites and associated clinical outcomes (Statement_40).

There were also calls for more flexible in screening guidelines, including proposals for risk-based screening and encouraging ‘on-demand’ online screening, though no alternatives commanded strong support (Statements_48/51). Table 2.2 presents illustrative statements.

#### 2.3 Management and treatment of asymptomatic cases of chlamydia and gonorrhoea in GBMSM

There were mixed views about whether to treat asymptomatic CT/NG infections, with nothing nearing consensus (Table 2.3). Some statements with mixed support argued for prompt, universal treatment (Statement_26/33/47), with one statement suggesting that antibiotic resistance in STIs is not a significant concern (Statement_22). In contrast, other statements argued against ‘needless’ treatment (Statements_18/35/52), stating that asymptomatic screening leads to overtreatment and increases the risk of AMR (Statements_1/9/83), that asymptomatic CT/NG infections do not pose harm to gay men (Statements_8/85), and that these infections will clear naturally over time (Statements_7/23/43). One statement suggested that we should consider NG/CT as commensals in asymptomatic individuals (Statement_85).

**Table 3:**
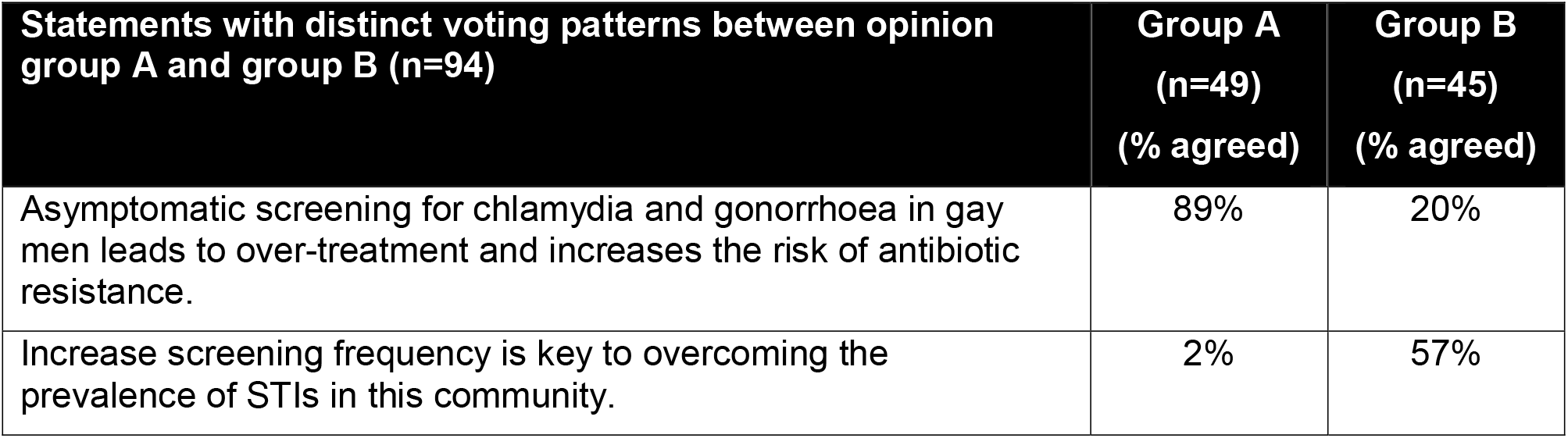
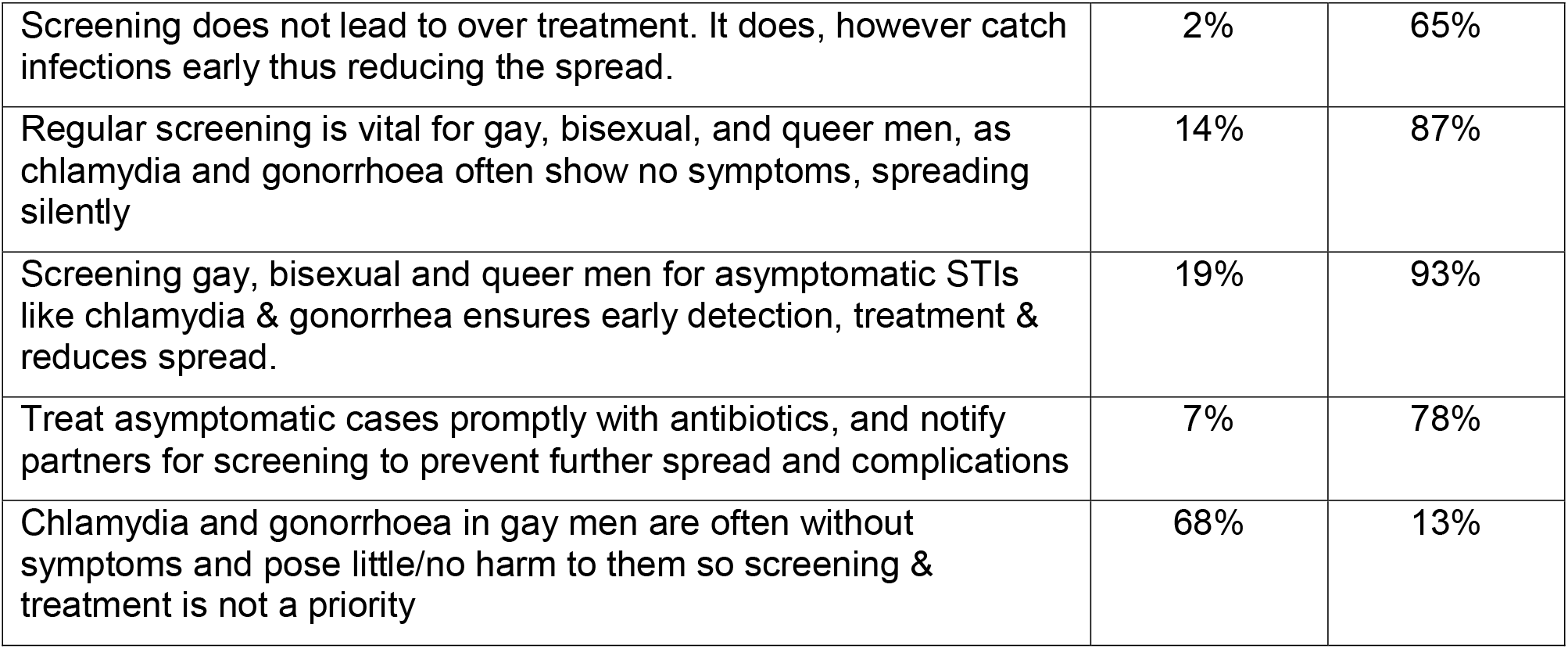
Voting patterns by opinion group A and B.

#### 2.4 Positive externalities and managing unintended consequences

Statements raised important knock-on effects (both positive and negative) of changing screening guidelines for CT/NG on other STIs, for other population groups, and for resource allocation, as well as strategies to mitigate these impacts (Table 2.4). In near unanimous agreement, participants concurred that it would be important to continue frequent screening for HIV and syphilis in the face of changes to other STI screening guidelines (Statement_76).

However, there were mixed views on whether reduced CT/NG screening would negatively affect screening for other STIs (Statements_2/34).

Numerous statements expressed significant concern about transmission across sexual networks, namely to heterosexual women, for whom there could be significant reproductive health consequences (Statements_3/11/16/56/57/64/66). There was mixed to moderate agreement that women should continue to be screened (and treated) for CT (Statement_56/64/66). While there were mixed to moderate agreement that bisexual men could infect women (Statements_3/16), there was near universal agreement that we need to avoid framing bisexual men as vectors of disease (Statement_90). Another statement raised the possibility that screening policies based on sexuality might exclude trans-women (but had low agreement) (Statement_89).

A range of statements highlighted the cost burden associated with frequent asymptomatic screening for CT/NG (Statements_13/24/25/36/37/50/59/79/80/81/84), with moderate agreement that reduced screening would produce significant cost-savings for HIV PrEP programs (Statement_14) and that resources could be more productively used to manage symptomatic patients (Statement_79). Other proposals for redirecting cost-savings (e.g. free HIV PrEP, controlling the syphilis epidemic, women-focused programs) had mixed support (Statements_24/25/36/59/81). There were mixed views as to whether reducing screening frequency would be perceived primarily as a cost-cutting exercise by community (Statement_78) or result in reduced funding for online screening services (Statement_10).

Community engagement and public health messaging was deemed essential for generating the traction needed to support changes to screening guidelines, and had strong support (Statements_58/60/68). While there was moderate agreement that asymptomatic screening provides a sense of safety and reassurance to GBMSM (Statements_20/77), there were mixed views about whether changes to screening recommendations would confuse community members and contradict decades of messaging about being a good sexual citizen (Statements_10/55/87).

### 3. Opinion groups

*Polis* algorithms categorized 94/99 participants into two ‘opinion groups’. Table 3 presents statements that exemplify voting differences between members of opinion group A and B.

Participants in opinion group A were more likely to believe that frequent screening does not reduce prevalence but leads to over-treatment and AMR risk in GBMSM for whom asymptomatic infections do not pose harm, while group B were more likely to hold opposite views across these fronts. Comparing the demographic profile of group A and B, we found that Group A had a higher proportion of participants from the UK and Europe while group B included a higher proportion from North America and low- and middle-income countries (<0.05). No other differences were found (Appendix 5).

## Discussion

The crowdsourcing platform, *Polis*, provided a useful tool for gauging the level of support for changing guidelines on CT/NG screening frequency in GBMSM. On the one hand, the results showed moderate support for decreasing screening frequency to 6-monthly, with support largely coalescing around resource savings, particularly for HIV PrEP programs. At the same time, there was a large divide in expert opinion regarding on what basis (i.e. clinical and epidemiological evidence) changes in screening frequency should be recommended and how asymptomatic cases should be managed. As shown by the voting patterns across opinion groups, those in favour of reduced screening differed from those opposed on three key fronts: (1) whether frequent asymptomatic screening plays a significant role in reducing community prevalence; (2) whether asymptomatic infections pose clinical harm and necessitate treatment; and, (3) whether frequent asymptomatic infections in GBMSM contributes to overtreatment and AMR. These findings point to important underlying views shaping expert opinion and that will need to be addressed in order to achieve greater consensus. Arguments around resource efficiency may provide a starting point for building common ground.

Our global exercise suggests confusion and/or lack of familiarity with emerging evidence likely contributed to low agreement. For example, nearly half of participants (inaccurately) agreed that frequent CT/NG screening reduces community prevalence, with many calling for RCTs to inform decision-making, despite the availability of real-world studies, including Gonoscreen^9^ and EZI-PREP.^27,28^ Further, there was significant uncertainty regarding natural clearance, as suggested by the significant proportion of participants who ‘passed’ on related statements (rather than vote for or against), contributing to a lack of consensus on how to manage asymptomatic cases.

One of the main arguments for reducing screening is to decrease the risk of AMR arising from the frequent antibiotic use, with estimates showing antibiotic consumption can be halved if screening among GBMSM is reduced.^29^ In addition to NG being no longer susceptible to the only remaining first-line antibiotic (ceftriaxone), increased NG AMR contributes to cross-resistance in non-STI related infections.^30^ Despite being a significant public health threat, our *Polis* exercise found mixed beliefs by the professional community on whether frequent screening contributes to overtreatment and AMR. While further education on AMR is certainly needed, differences in voting patterns by UK/European participants versus North Americans (as observed in the opinion groups) may point to political and cultural differences in the way that individual risk versus population-level risks are collectively prioritized. Opinions based on values may be harder to shift.

Given the increasing incidence of global syphilis (including congenital syphilis), and strong agreement among study participants that screening for women and for HIV and syphilis must not be impacted by reduced CT/NG screening among GBMSM, strategies will be needed to ensure high vigilance in detecting syphilis, especially among men who have sexual contact with women. Further, community engagement and future research is needed to understand effective ways of conveying nuanced and differentiated screening messages for different population groups without creating confusion or resistance. New messages and ways of understanding what it means to be a ‘good sexual citizen’ will be needed to shift the ingrained ‘test and treat’ approach.

Using *Polis* allowed us to reap benefits offered by both qualitive and quantitative methods. It allowed participants to present arguments and concerns (in their own words) that they considered most salient to the debate, while also generating structured, and scalable quantitative data. However, participant-generated statements which were phrased unclearly may have contributed to low agreement. Our study supported global participation through an asynchronous, interactive, and anonymous platform. However, our recruitment methods may have failed to reach important professional networks, including in the Global South.

## Conclusion

Frequent asymptomatic screening for CT/NG in GBMSM consumes significant resources for uncertain benefit in this population, while contributing to AMR. While our global crowdsourcing exercise was intended to identify areas of consensus, it instead illustrated a large divide in expert opinion on many fronts. The findings suggest that many in the sexual health field may not be fully versed with the evidence that is needed to support a well-informed dialogue on whether to challenge or retain the current ‘test and treat’ paradigm. As such, agreement leaned towards an incrementalist approach to reducing screening frequency rather than a full-scale change to screening and treatment of asymptomatic CT/NG in GBMSM.

## Data Availability

All data produced in the present study are available upon reasonable request to the authors

## Acknowledgements

We would like to acknowledge Eloise Williams, Matthew Golden, Matthew Vaughan, Nicholas Medland, Janneke Heijne, Michael Kwag, who participated in a panel discussion of these results and asymptomatic screening more generally at the 2024 IUSTI World Congress.

