## Supplementary appendices for "Should we screen less frequently for chlamydia and gonorrhoea in gay and bisexual men who have sex with men? Findings from a global crowdsourcing exercise with experts"

Appendix 1: Seed Statements

| ID | Seed statement |
| --- | --- |
| 0 | We don’t need to treat asymptomatic chlamydia or gonorrhoea because our body will eliminate the infection over time |
| 1 | The main benefit of screening is to benefit the health of individuals rather than preventing transmission into the community. |
| 2 | It’s too hard to create clear messaging around the need to test regularly for HIV and syphilis, but not for gonorrhoea and chlamydia |
| 3 | Changing guidelines for asymptomatic testing will water down the efforts of the queer community to raise awareness about the importance of testing |
| 4 | Bisexual men who don’t get tested regularly risk putting heterosexual women at risk of chlamydia and gonorrhoea |
| 5 | Reducing the frequency of screening for chlamydia and gonorrhoea in gay men will have negative knock-on effects on the frequency of screening for HIV and syphilis |
| 6 | Testing and treating STIs has not worked to reduced community prevalence and associated disease for STIs |
| 7 | Asymptomatic screening for chlamydia and gonorrhoea in gay men leads to over-treatment and increases the risk of antibiotic resistance |

Appendix 2: Rejected Statements

| **Submitted statement** | **Reason for rejecting** |
| --- | --- |
| Multiplex PCR platforms produce positive test results without clinical significance and are thus uneconomical and undesirable | Off topic |
| Multiplex PCR platforms produce unwanted positive test results without clinical significance and thus hinder clinicians | Off topic |
| The wave of laws and regulations to ban the advocacy and funding of LGBT people would not help the fight of HIV in Africa | Off topic |
| Decriminalizing of sex work would go a long way to help the fight for all sexually transmitted infections. | Off topic |
| Asymptomatic screening in MSM can be reduced to 6 monthly | Duplicate |
| More funds should be made available to control syphilis and independently evidence-based criteria for screening for bacterial STIs | Duplicate |

Appendix 3: All included statements and votes

**STATEMENT**

0

Asymptomatic screening for

chlamydia and gonorrhoea in gay

men leads to over-treatment and

increases the risk of antibiotic

resistance

60

%

%

17

%

22

(

81

)

89

%

2

%

8

%

(

47

)

20

%

38

%

41

%

(

34

)

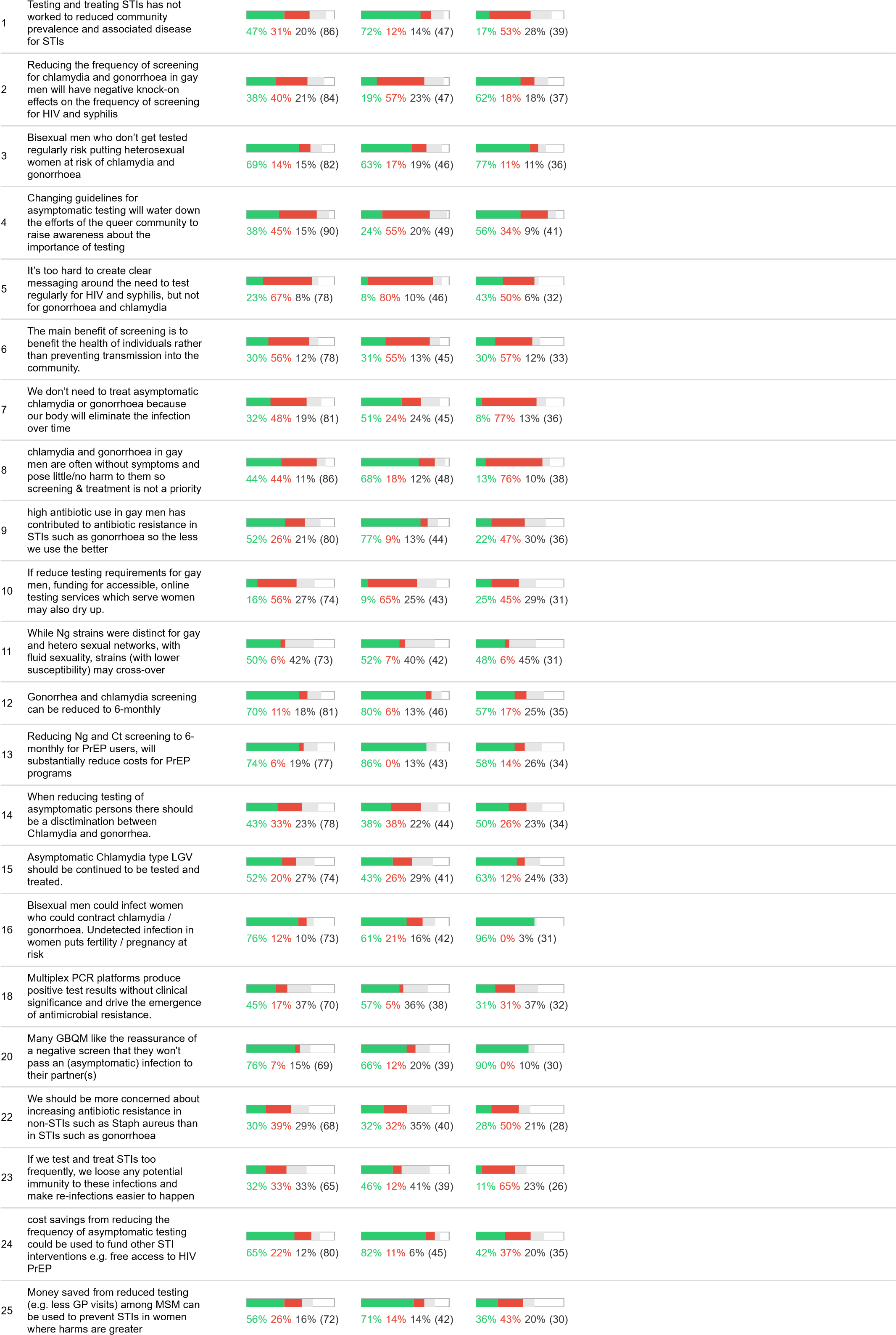

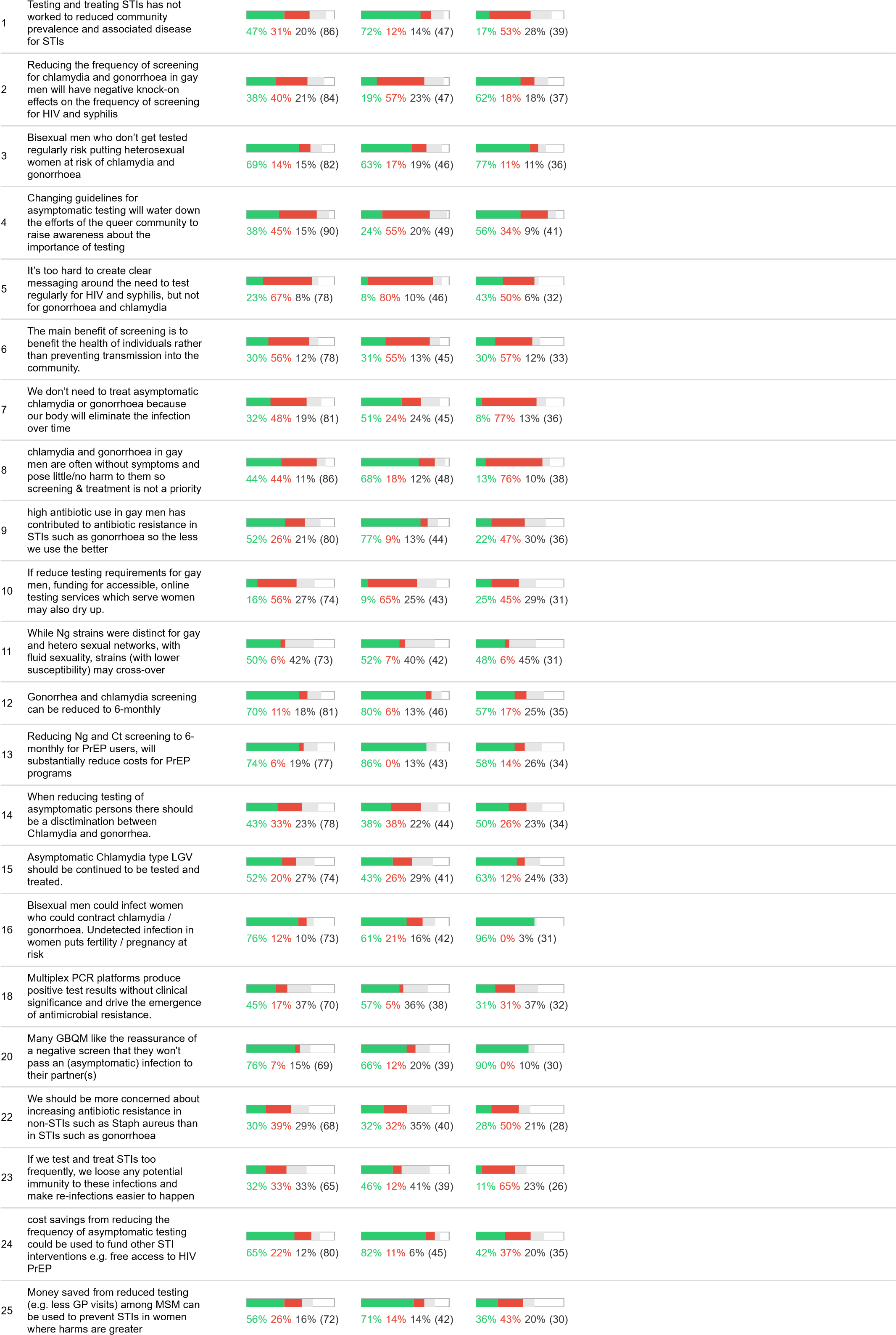

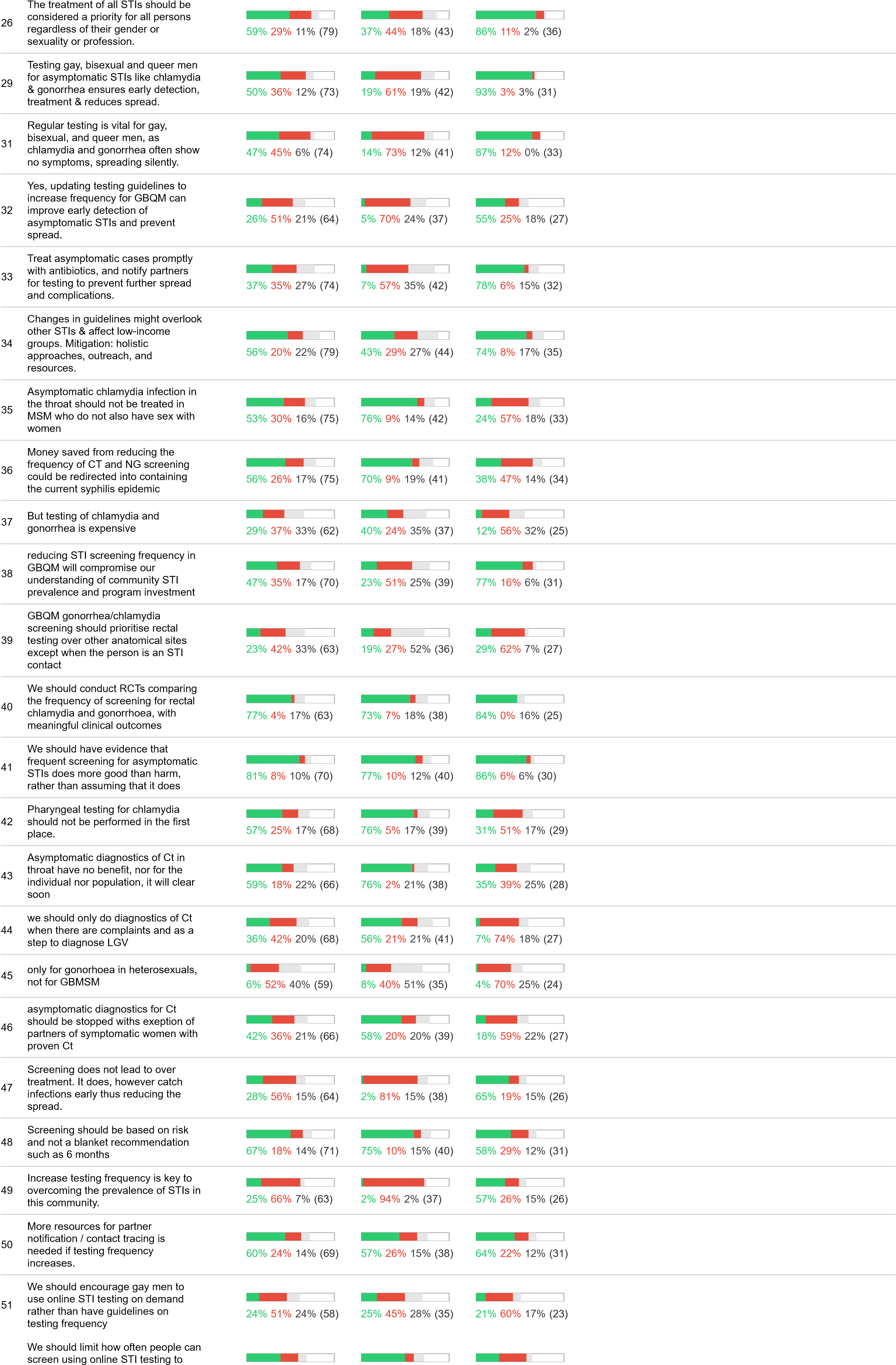

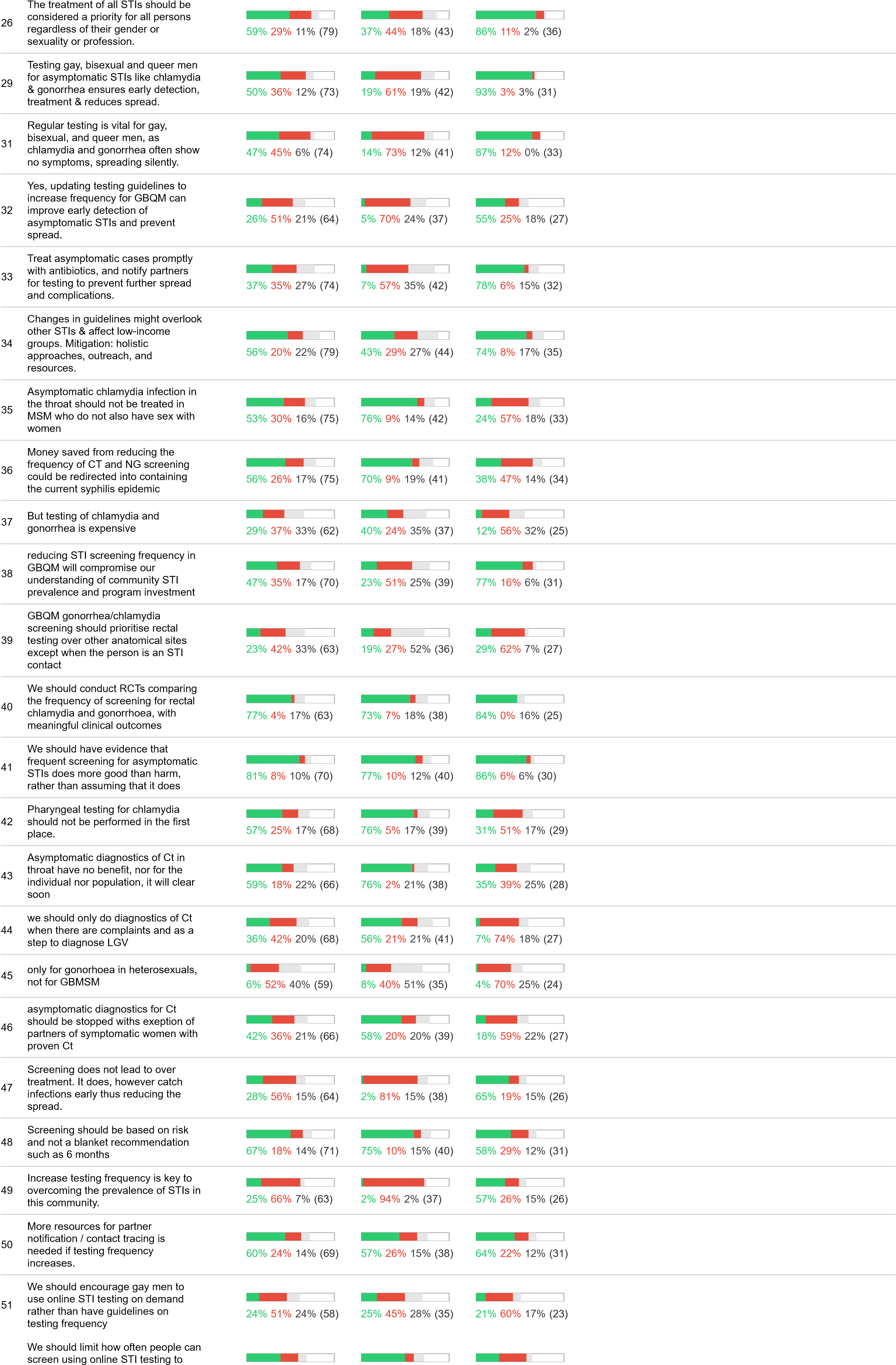

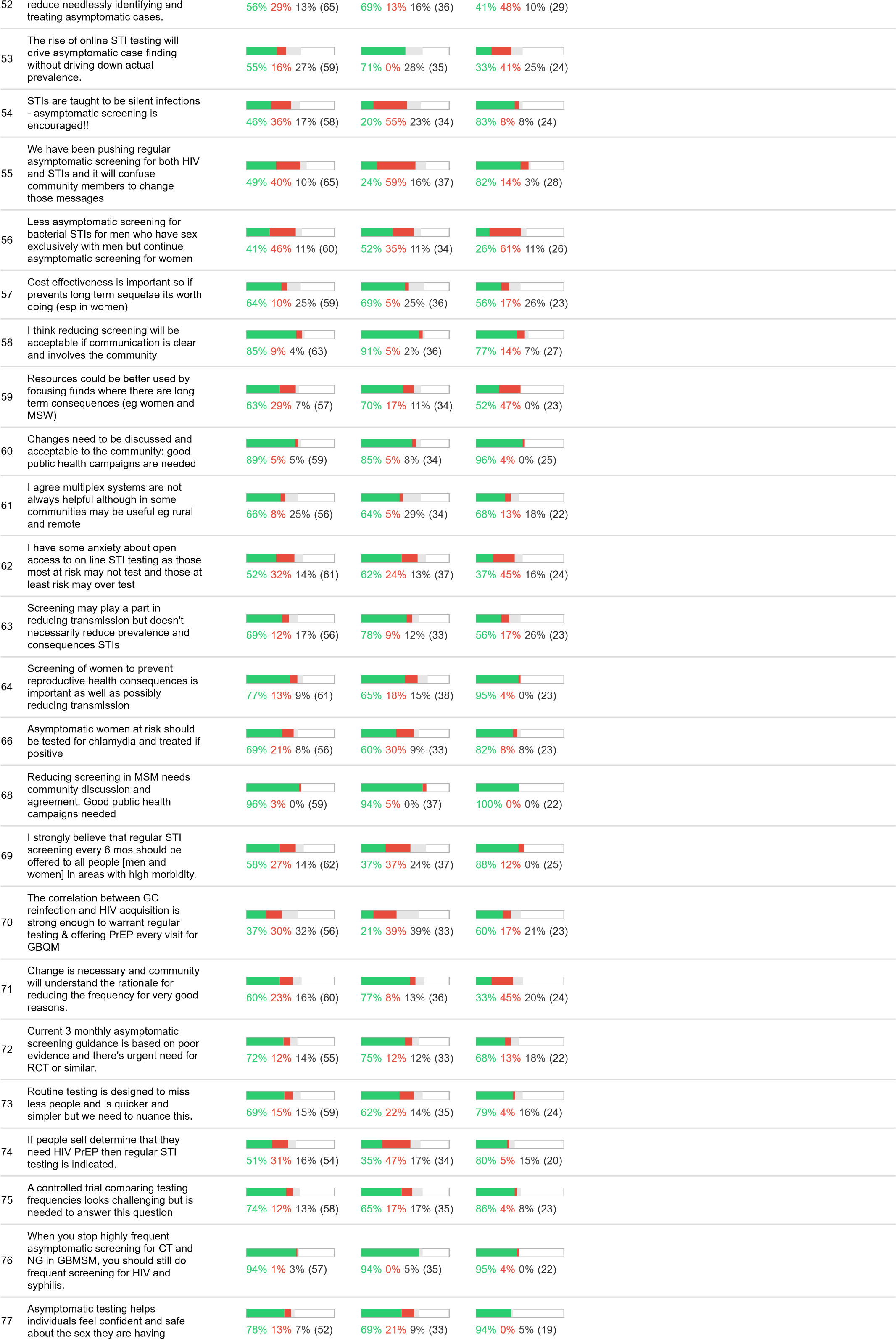

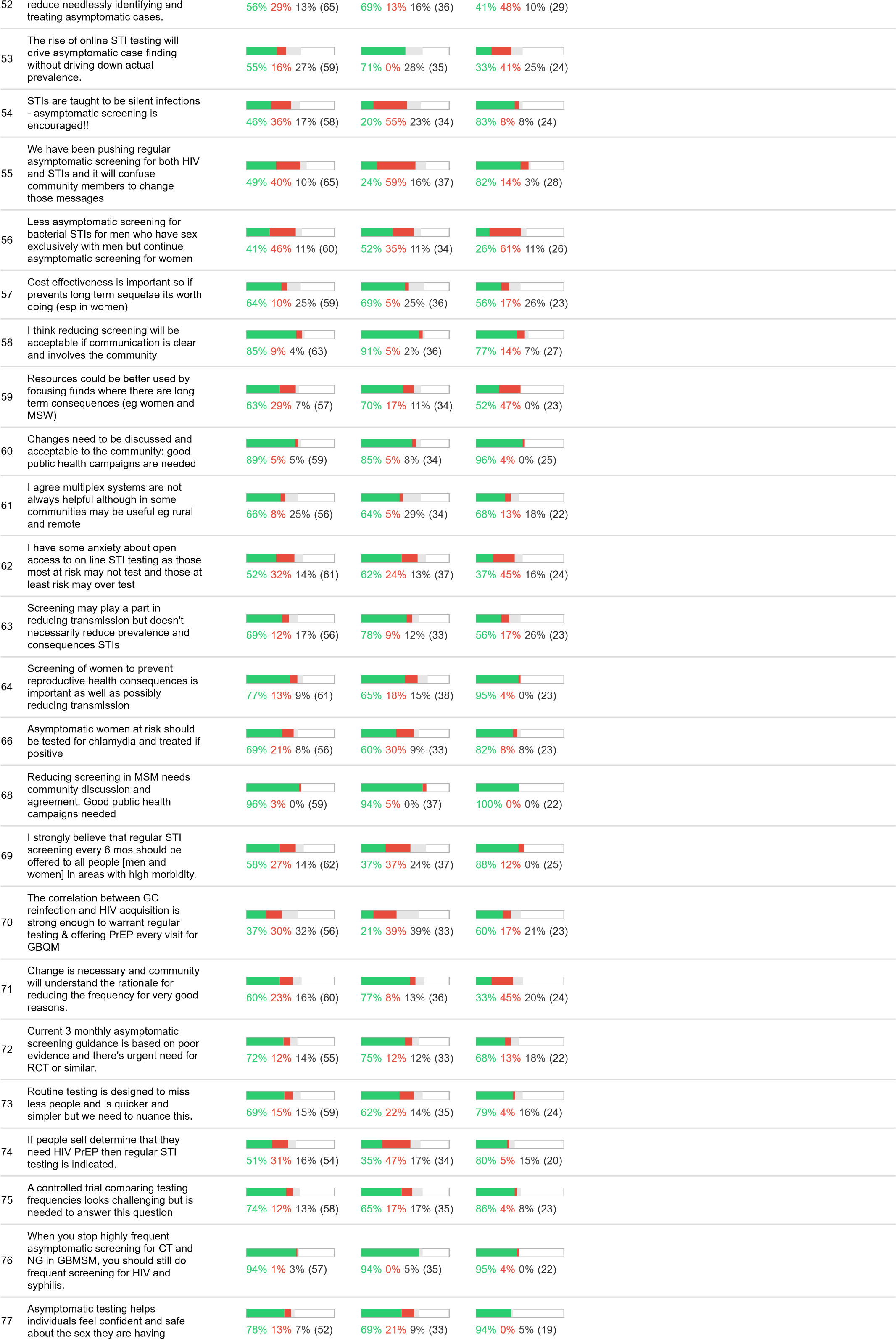

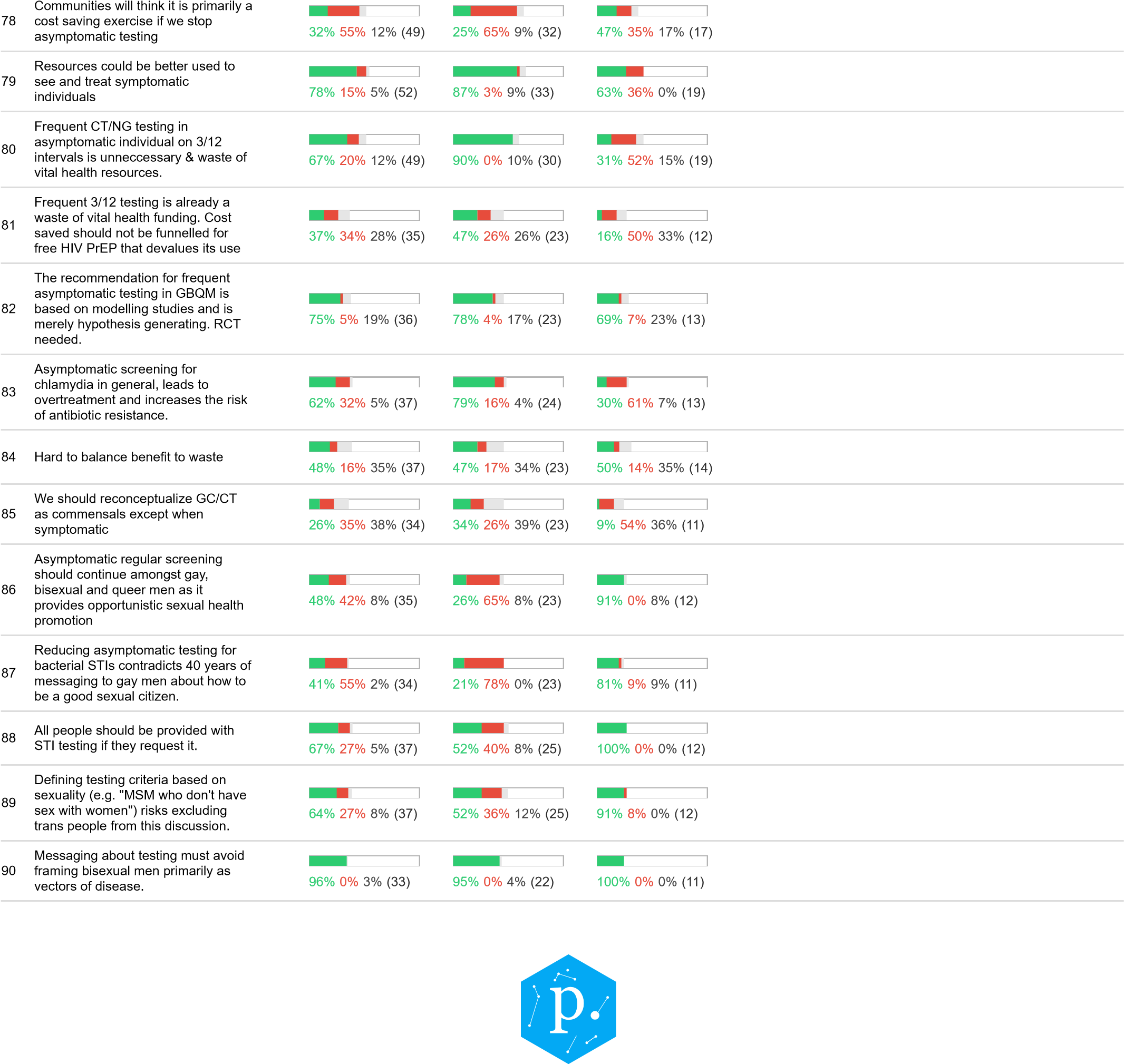

Appendix 4: Registration survey

Start of Block: Survey information

**Should we change recommendations on the frequency of asymptomatic testing for Chlamydia and Gonorrhoea and in Gay, Bisexual and Queer Men?**

Thanks for your interest in taking part of the conversation on asymptomatic testing.

**Complete the form below to register. A web-link will be emailed to you to join the conversation on Polis.**

Polis is a real-time system for understanding what large groups of people think. You will be shown statements that other people have written - vote either ‘agree’, ‘disagree’ or ‘pass’. You can write a new statement if you like that others can vote on.  Your votes and comments will be anonymous. As time goes on, **you’ll be able to see a map of the opinion groups that emerge.**

Please check into Polis twice a week to vote on new opinions. We'll remind you.

View the plain language statement from the University of Melbourne and University of Toronto for more information

| Page Break |
| --- |

Consent 1 I consent to participating in this Polis discussion about asymptomatic testing for chlamydia and gonorrhoea in gay, bisexual and queer men.

- Yes, I consent (1)

Q1 Click to show you're not a robot

End of Block: Survey information

Start of Block: Registration form and consent

Q2 First name

________________________________________________________________

Q3 Email address (we need this to send you a unique weblink to participate in the online Polis conversation)

________________________________________________________________

Q4 What is your gender?

- Man (1)
- Woman (2)
- Non-binary (3)
- Self-identify (4) __________________________________________________
- Prefer not to say (5)

Q5 Do you identify as gay, bisexual or queer?

- Yes (1)
- No (2)
- Prefer not to say (3)

Q6 What is your main professional affiliation?

▼ Nursing (1)

Other Allied Health (2)

Diagnostics (3)

General Practice Physician (4)

Sexual Health Physician (5)

Medical Specialist – Infectious diseases (6)

Medical Specialist – Other (7)

Pathology/Laboratory (8)

Pharmacy (9)

Social Work/Counselling (10)

Education (11)

Health Promotion (12)

Legal (13)

Policy (14)

Research – Basic (15)

Research – Clinical (16)

Research – Epidemiology (17)

Research – Public Health (18)

Research – Social (19)

Sexology (20)

Women’s Health (21)

Other (please describe) (22)

Q7 What is your Primary Work setting?

▼ University/Research Institute (1)

Public Healthcare (2)

Private Healthcare (3)

Government (local, state or national) (4)

Non-governmental or other non-profit organization (5)

Industry (6)

Other (7)

Q8 Are you a graduate student (e.g. Master’s/doctoral) or trainee?

- Yes (1)
- No (2)

| Page Break |
| --- |

Q9 Where is the organization that you (primarily) work for located? **Write the name of the country in the open-text field below each option.**

- Africa (1) __________________________________________________
- Asia (Central) (4) __________________________________________________
- Asia (East/Southeast/South) (2) __________________________________________________
- Latin America and Caribbean (6) __________________________________________________
- North America (10) __________________________________________________
- Oceania (5) __________________________________________________
- UK and Europe (3) __________________________________________________

Q10 How would you rate your current level of knowledge regarding treatment and management of gonorrhoea and chlamydia?

- None	(1)
- Limited	(2)
- Moderate	(3)
- Good	(4)
- Expert	(5)

Q11 How would you rate your current level of knowledge regarding STI-related drug resistance?

- None	(1)
- Limited	(2)
- Moderate	(3)
- Good	(4)
- Expert	(5)

Q12 To what extent to you agree with these statements?

|  | Strongly agree (1) | Generally agree (2) | Neutral (3) | Generally disagree (4) | Strongly disagree (5) | Unsure (6) |
| --- | --- | --- | --- | --- | --- | --- |
| Antibiotic resistance is a serious threat to global public health (1) |  |  |  |  |  |  |
| I am not concerned with antibiotic resistance in my country at the present time (2) |  |  |  |  |  |  |

Contact Q Are you interested in participating in an interview on this topic?

- Yes (1)
- No (2)

| Page Break |
| --- |

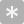

Q13 Answer this question to make sure you're not a robot. We will present these results at the 2024 International Union against STIs World Congress. What is the acronym for this conference?

- Australia Forum (1)
- IUSTI (2)
- G8 (3)
- Global Health Conference (4)
- Annual STI Diagnostics Conference (5)

**Go to 'Next Page' to Submit**

Background on the topic: In high-income countries, STI screening guidelines for men who have sex with men typically recommend screening every 3-6 months. Most chlamydia and gonorrhoea infections detected in men who have sex with men are asymptomatic. There is increasing debate about the harms and benefits of frequent, asymptomatic testing for gonorrhoea and chlamydia in this population. Some evidence suggests that frequent screening does not substantially reduce the prevalence of gonorrhoea and chlamydia in the population, but contributes to increased antibiotic use and antibiotic resistance in the population. At present, gonorrhoea is already resistant to multiple classes of antibiotics. On the other hand, the importance of regular STI testing, for a range of STIs including HIV and syphilis, has been an important message promoted in the gay, bisexual and queer men community to keep individuals safe. The risk of HIV transmission may be higher in individuals with asymptomatic infections; however, PrEP is highly efficacious even in the presence of chlamydia and gonorrhoea infections. Further, for men who have sex with men and women, transmission of chlamydia and gonorrhoea to women can have serious consequences, including leading to pelvic inflammatory disease and fertility issues.

End of Block: Registration form and consent

Appendix 5: Characteristics of participants in opinion group A and B

|  |  | Opinion Group A |  | Opinion Group B |  | *p*-value |
| --- | --- | --- | --- | --- | --- | --- |
|  |  | n=49 | % | n=45 | % |  |
| Gender | Woman | 34 | 69·4 | 24 | 53·3 | 0·32 |
|  | Man | 14 | 28·6 | 19 | 42·2 |  |
|  | Non-binary | 1 | 2·0 | 2 | 4·4 |  |
| LGBTIQ+ | Yes | 21 | 42·9 | 18 | 40·0 | 0·71 |
| Profession | Clinician | 19 | 38·8 | 20 | 44·4 | 0·27 |
|  | Researcher | 20 | 40·8 | 15 | 33·3 |  |
|  | Education/social work/counselling/health promotion | 2 | 4·1 | 5 | 11·1 |  |
|  | Other | 8 | 16·3 | 5 | 11·1 |  |
| Primary work setting | Government | 13 | 26·5 | 8 | 17·8 | 0·22 |
|  | NGO | 3 | 6·1 | 5 | 11·1 |  |
|  | Private healthcare | 0 | 0 | 2 | 4·4 |  |
|  | Public Healthcare | 19 | 38·8 | 12 | 26·7 |  |
|  | University/Research Institute | 14 | 28·6 | 16 | 35·6 |  |
|  | Other | 0 | 0 | 2 | 4·4 |  |
| Region | Africa/Asia/Latin America | 0 | 0 | 7 | 15·6 | <0·001 |
|  | North America | 6 | 12·2 | 15 | 33·3 |  |
|  | Oceania | 21 | 42·9 | 17 | 37·8 |  |
|  | UK and Europe | 22 | 44·9 | 6 | 13·3 |  |
| Knowledge management of NG and CT | Expert | 22 | 44·9 | 14 | 31·1 | 0·64 |
|  | Good | 19 | 38·8 | 21 | 46·7 |  |
|  | Moderate | 7 | 14·3 | 8 | 17·8 |  |
|  | Limited | 1 | 2·0 | 1 | 2·2 |  |
|  | None | 0 | 0·0 | 1 | 2·2 |  |
| Knowledge of AMR | Expert | 12 | 24·5 | 12 | 26·7 | 0·30 |
|  | Good | 25 | 51·0 | 16 | 35·6 |  |
|  | Moderate | 10 | 20·4 | 10 | 22·2 |  |
|  | Limited | 2 | 4·1 | 6 | 13·3 |  |
|  | None | 0 | 0·0 | 1 | 2·2 |  |
